# Genetic Polymorphisms Associated with Adverse Pregnancy Outcomes in Nulliparas

**DOI:** 10.1101/2022.02.28.22271641

**Authors:** Rafael F. Guerrero, Raiyan R. Khan, Ronald J. Wapner, Matthew W. Hahn, Anita Raja, Ansaf Salleb-Aouissi, William A. Grobman, Hyagriv Simhan, Robert Silver, Judith H. Chung, Uma M. Reddy, Predrag Radivojac, Itsik Pe’er, David M. Haas

## Abstract

**Background:** Adverse pregnancy outcomes (APOs) affect a large proportion of pregnancies and represent an important cause of morbidity and mortality worldwide. Yet, the pathophysiology of APOs is poorly understood, limiting our ability to prevent and treat these conditions.

**Objective:** To search for genetic risk markers for four APOs, we performed genome-wide association studies (GWAS) for preterm birth, preeclampsia, gestational diabetes, and pregnancy loss.

**Study Design:** A total of 9,757 nulliparas from the nuMoM2b study were genotyped. We clustered participants by their genetic ancestry and focused our analyses on the three sub-cohorts with the largest sample sizes: European (EUR, n=6,082), African (AFR, n=1,425), and American (AMR, n=846). Association tests were carried out separately for each sub-cohort and brought together via meta-analysis. Four APOs were tested by GWAS: preeclampsia (n=7,909), gestational length (n=4,781), gestational diabetes (n=7,617), and pregnancy loss (n=7,809). Using the results of the genome-wide associations for each APO, SNP-based heritability of these traits was inferred using LDscore. Putative regulatory effects were inferred by transcriptome-wide association analysis.

**Results:** Two variants were significantly associated with pregnancy loss (rs62021480: OR = 3.29, *P* = 7.83×10^−11^, and rs142795512: OR = 4.72, *P* = 9.64×10^−9^), implicating genes *TRMU* and *RGMA* in this APO. An intronic variant was significantly associated with gestational length (rs73842644: beta = -0.667, *P* = 4.9×10^−8^). Three loci were significantly associated with gestational diabetes (rs72956265: OR = 3.09, *P* = 2.98×10^−8^, rs10890563: OR = 1.88, *P* = 3.53×10^−8^, rs117689036: OR = 3.15, *P* = 1.46×10^−8^), located on or near *ZBTB20, GUCY1A2*, and *MDGA2*, respectively. Several loci previously correlated with preterm birth (in genes *WNT4, EBF1, PER3, IL10*, and *ADCY5*), gestational diabetes (in *TCF7L2*), and preeclampsia (in *MTHFR*) were found to be associated with these outcomes in our cohort as well.

**Conclusion:** Our study identified genetic associations with gestational diabetes, pregnancy loss, and gestational length. We also confirm correlations of several previously identified variants with these APOs.

**Disclosure Statement:** The authors declare no conflict of interest

**Source of financial support:** Precision Health Initiative of Indiana University, National Institutes of Health award R01HD101246 to DMH and PR. Cooperative agreement funding from the National Heart, Lung, and Blood Institute and the Eunice Kennedy Shriver National Institute of Child Health and Human Development: grant U10-HL119991 to RTI International; grant U10-HL119989 to Case Western Reserve University; grants U10-HL120034 and R01LM013327 to Columbia University; grant U10-HL119990 to Indiana University; grant U10-HL120006 to the University of Pittsburgh; grant U10-HL119992 to Northwestern University; grant U10-HL120019 to the University of California, Irvine; grant U10-HL119993 to University of Pennsylvania; and grant U10-HL120018 to the University of Utah. National Center for Research Resources and the National Center for Advancing Translational Sciences, National Institutes of Health to Clinical and Translational Science Institutes at Indiana University (grant UL1TR001108) and University of California, Irvine (grant UL1TR000153).

## Introduction

Adverse pregnancy outcomes (APOs) are a serious threat to the health of pregnant persons and children. APOs affect a significant fraction of pregnancies across the globe and are among the leading causes of morbidity and mortality worldwide^1^. Among the most common APOs are preterm birth (which occurs in over 10% of pregnancies in the United States (US)^2^), preeclampsia (which develops in 5-10% of pregnancies^3^), gestational diabetes (occurring in roughly 6% of pregnancies in the US^4^), and pregnancy loss (estimated to occur in about 20% of pregnancies in the US^5^). APOs are also highly correlated with future disease in birthing parents. For example, gestational diabetes carries a lifetime 50% risk of type 2 diabetes (T2D) in the mother^6^, while preeclampsia might lead to a 2-3 fold increase of cardiovascular disease later in life^7^. Yet, the factors driving these diseases remain poorly understood, hindering efforts in prevention and treatment.

To better understand the mechanisms and improve the prediction of APOs in nulliparous individuals, the Nulliparous Pregnancy Outcomes Study: Monitoring Mothers-to-Be (nuMoM2b) consortium recruited and prospectively followed a large cohort of nulliparous people beginning in their first trimester of pregnancy. Participants underwent several assessments over the course of their pregnancies, resulting in a comprehensive profile that included biospecimens, clinical measurements, ultrasounds, behavior (through interviews and questionnaires), physical activity assessment, and dietary content.

By precisely characterizing different aspects of over 10,000 pregnancies, the nuMoM2b cohort has already yielded valuable insights into the factors that contribute to APOs^8,9,10^. Additionally, the availability of biospecimens provides a unique opportunity to study the genetic underpinnings of APOs. The objective of this study was to genotype the nuMoM2b cohort and to search genome-wide for variants associated with four APOs: gestational length (as a proxy for preterm birth), preeclampsia, gestational diabetes mellitus (GDM), and pregnancy loss.

## Methods

### Participants

The participants of the analysis were enrolled in the nuMoM2b cohort (https://www.nichd.nih.gov/research/supported/nuMoM2b), a longitudinal, multiethnic cohort study of nulliparous individuals. All participating centers obtained approval by the local Institutional Review Boards (IRBs) of their corresponding recruitment institutions, with further details on data collection having been previously described^11,12^. The study enrolled 10,038 nulliparous people from the first trimester of their pregnancy to participate in three study visits during pregnancy. Collection of health status and biomarkers were conducted at regular intervals, and documentation of pregnancy outcomes was performed by chart abstraction using *a priori* definitions. The details of this process were described elsewhere^7^.

### Phenotype definitions

Gestational length: We opted to use a quantitative phenotype, gestational length, instead of a binary preterm/full term outcome to gain additional information and statistical power by using a more granular phenotype. Gestational length was determined from an estimated due date established by a first-trimester ultrasound crown-rump length measurement and was recorded in weeks^12^. Preterm birth was defined as any live birth that occurred before 37 weeks gestational age. Cases of stillbirth, fetal demise, elective termination, and indicated termination were all excluded from this phenotype group (Table 1).

**Table 1.** Number of subjects in the nuMoM2b cohort used for the GWAS. Each row represents the number of subjects remaining after applying the step. Bolded numbers represent the final number of subjects used for each separate GWAS (GDM: gestational diabetes, GL: gestational length, PEC: preeclampsia, PL: pregnancy loss).

|  | African Ancestry |  |  |  | Admixed American Ancestry |  |  |  | European Ancestry |  |  |  |
| --- | --- | --- | --- | --- | --- | --- | --- | --- | --- | --- | --- | --- |
| **Initial** | 1425 |  |  |  | 846 |  |  |  | 6082 |  |  |  |
| **Preprocessing** | 1384 |  |  |  | 811 |  |  |  | 5896 |  |  |  |
| **After removal of related Subjects** | 1374 |  |  |  | 811 |  |  |  | 5891 |  |  |  |
| **GWAS** | GDM | GL | PEC | PL | GDM | GL | PEC | PL | GDM | GL | PEC | PL |
| **Available Phenotypes** | **1258** | 1355 | **1355** | **1308** | **754** | 775 | **775** | **775** | **5605** | 5726 | **5779** | **5726** |
| **Live and Spontaneous** | - | **770** | - | - | - | **512** | - | - | - | **3499** | - | - |

Preeclampsia: Cases include diagnosis of preeclampsia (with and without severe features), eclampsia, and chronic hypertension with superimposed preeclampsia. A detailed description of nuMoM2b study definitions of hypertensive disorders of pregnancy was published in the supplement to the paper by Facco et al^13^. All other individuals were treated as controls.

Gestational diabetes (GDM): GDM was diagnosed through clinical evaluation, from fasting blood sugar, sequential 1-hour glucose challenge test followed by a 3-hour glucose tolerance test (GTT), or a single step 2-hour 75-gram GTT^13^. We excluded individuals diagnosed with pregestational diabetes from GDM analyses, and all other individuals were treated as controls. Pregnancy loss: All subjects who had a pregnancy loss, regardless of gestational age, were considered as cases (Table 1). Individuals who underwent termination of pregnancy were excluded from the pregnancy loss analysis. Subjects who had a live birth were treated as controls.

### Genotyping

The genotyped cohort comprised 9,757 participants who had adequate samples and agreed to be genotyped. DNA extractions from whole blood were carried out on a QIAsymphony instrument (from Qiagen; extraction kit DSP DNA Midi Kit #937355, protocol Blood_1000_V7_DSP) at the Center for Genomics and Bioinformatics (Indiana University, Bloomington). Genotyping was completed at the Van Andel Institute (Grand Rapids, MI, USA) using the Infinium Multi-Ethnic Global D2 BeadChip (Illumina, Miami, USA). We imposed standard filters for quality control of loci at this stage (cluster separation < 0.3, AA R Mean < 0.2, AB R Mean < 0.2, BB R Mean <0.2, 10% GC < 0.3) using GenomeStudio v2.4 (Illumina). Genotype calls (in GCT format) for the 1,748,280 loci that passed initial quality control were made with Beeline autoconvert (Ilumina).

### Inclusion criteria, QC pipeline, Ancestry determination, and imputation

We performed preprocessing to determine any sex inconsistencies, autosome missingness (>5%), and contamination (more than 25 related subjects within the same dataset). Given the highly heterogeneous data set, we used the KING-Robust algorithm, a pairwise kinship estimator for GWAS that is robust to the presence of unknown population substructure^14^. We inferred kinship estimates between all pairs of subjects, randomly removing one subject from the pairs of subjects with first- or second-degree relatedness such that we minimized the number of subjects removed. The remaining samples were filtered to remove SNPs with a minor allele frequency (MAF) less than 0.01, genotyping rate <95%, and a Hardy-Weinberg Equilibrium (HWE) *P* < 5×10^−6^.

We determined the ancestry of each subject using SNPweights v.2.1 and leveraging approximately 40,000 ancestry informative markers available from the 1000 Genomes Consortium^15^. Applying a probability cut-off of >50%, the samples were clustered into five ancestry groups concordant with the 1000 Genomes Consortium^15^ super-populations: African (AFR, n=1425), American (AMR; n=846), East Asian (EAS; n=323), European (EUR, n=6082), and South Asian (SAS; n=112). We observed a large fraction of highly admixed individuals, thus we also established a sixth group (ADM, n=891) of subjects who have none of the five ancestry group percentages reaching >50% (Supplemental Figure S1). The heterogeneous genetic ancestry of this cohort would result in high levels of genomic inflation in a single GWAS analysis. To reduce this confounding effect, we clustered the cohort by genetic ancestry for downstream analyses (Supplemental Figure S1, Supplemental Figure S2). Thus, from this point subjects were analyzed in separate groups according to their majority genetic ancestry when doing the filtering preprocessing and imputation.

Not all the ancestry groups contained enough subjects to power a GWAS study. Thus, we only proceeded with imputing the European, African, and American ancestry subjects. These subjects were phased with EAGLE and imputed by Minimac3^16^ using the TOPMED Imputation Server^17,18^. We used version R2 of the TOPMED panel, currently the largest panel of sequenced human genomes, and containing all the ancestries we observed in our cohort. Following imputation, we excluded SNPs with an *r*^2^ quality score <0.7, MAF <0.01, genotyping rate <95%, and a Hardy-Weinberg Equilibrium (HWE) *P* < 5×10^−6^ within each imputed group.

Population structure was determined by PLINK using pruned SNPs from the data (Linkage Disequilibrium pruning *r*^2^ < 0.1). The top ten principal components were computed for each of the three sub-cohorts (EUR, AFR, AMR). As maternal age has a non-linear effect on preterm birth, we transformed each subject’s age into the distance from the median percentile in all subjects.

### Genome-wide associations

Association testing was carried out using regression models implemented in PLINK v1.9^19^. The suggestive association threshold was *P* < 1×10^−5^ and the threshold for genome-wide significance was *P* < 5×10^−8^. The model was adjusted for each subject’s rank-transformed age and the first ten principal components from our population structure analysis. We used all ten computed principal components due to the high level of population substructure and genetic admixture present in the nuMoM2b data set. Any subjects with missing age or phenotype information were excluded from the analysis. Results from each sub-cohort were fixed-effect meta-analyzed using GWAMA^20^ and SNPs that did not appear in at least two of the three individual GWAS were removed. Result plots were displayed using R libraries. The SNPs of interest were further annotated with Variant Effect Predictor^21^ to identify their corresponding gene function. The model for gestational length was adjusted for rank-transformed age.

Using the results of the genome-wide associations for each APO, we inferred SNP-based heritability of these traits using LDscore^22^. We searched for phenotypes that had significant genetic correlations with the APOs using the LD-Hub database^23^. Finally, we scanned for putative regulatory effects by carrying out a transcriptome-wide association analysis (TWAS) as implemented in the Fusion software, and using the available expression reference weights for whole blood and adipose tissue^24^, as well as liver, pancreas, vagina, and uterus, and whole blood from the GTEx v7 multi-tissue RNA-seq data set. For TWAS, *P*-values were Bonferroni-corrected by the number of genes in each panel. Conditional expression analyses were performed using R scripts from the Fusion software.

## Results

### Pregnancy Loss

This trait showed the highest SNP-based heritability (0.25) of the four APOs studied. We observed two novel associations with pregnancy loss (Table 2): rs62021480 and rs142795512. Located on chromosome 15, rs62021480 encodes a synonymous variant on *RMGA* (Repulsive Guidance Molecule BMP Co-Receptor A), a gene that encodes a glycoprotein that guides developing axons and may act as a tumor suppressor. Rs142795512 is an intronic variant in the gene *TRMU* (TRNA 5-Methylaminomethyl-2-Thiouridylate Methyltransferase), which encodes a mitochondrial tRNA-modifying enzyme. Variants in *TRMU* have been linked to several disease phenotypes, including infantile liver failure^25^.

**Table 2.**
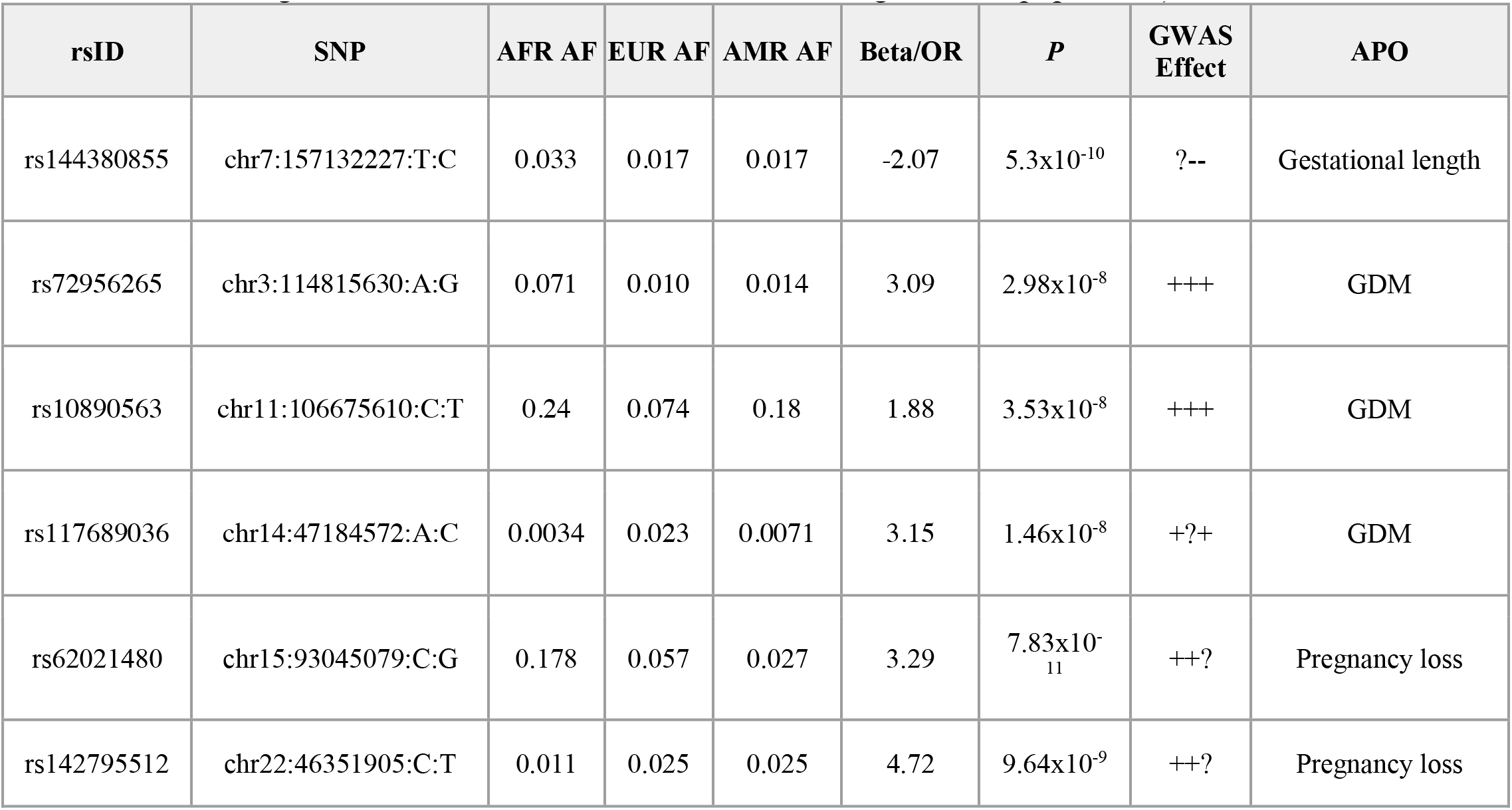
All genome-wide significant SNPs across the four GWAS analyses performed. The allele frequency at the population level (as reported on Gnomad^32^) is indicated for African ancestry (AFR AF), European ancestry (EUR AF), and Admixed American ancestry (AMR AF). Odds ratio (OR) is computed for the pregnancy loss (logistic regression) GWAS, while beta is computed for the gestational length (GWAS). “GWAS effect” reflects the direction of the odds ratio or beta of the SNP in each sub-cohort (EUR, AFR, AMR; + indicates positive effect, - indicates negative effect, ? indicates the SNP was missing from the population).

| rsID | SNP | AFR AF | EUR AF | AMR AF | Beta/OR | *P* | GWAS Effect | APO |
| --- | --- | --- | --- | --- | --- | --- | --- | --- |
| rs144380855 | chr7:157132227:T:C | 0.033 | 0.017 | 0.017 | -2.07 | 5.3x10<sup>-10</sup> | ?-- | Gestational length |
| rs72956265 | chr3:114815630:A:G | 0.071 | 0.010 | 0.014 | 3.09 | 2.98x10<sup>-8</sup> | +++ | GDM |
| rs10890563 | chr11:106675610:C:T | 0.24 | 0.074 | 0.18 | 1.88 | 3.53x10<sup>-8</sup> | +++ | GDM |
| rs117689036 | chr14:47184572:A:C | 0.0034 | 0.023 | 0.0071 | 3.15 | 1.46x10<sup>-8</sup> | +?+ | GDM |
| rs62021480 | chr15:93045079:C:G | 0.178 | 0.057 | 0.027 | 3.29 | 7.83x10<sup>-11</sup> | ++? | Pregnancy loss |
| rs142795512 | chr22:46351905:C:T | 0.011 | 0.025 | 0.025 | 4.72 | 9.64x10<sup>-9</sup> | ++? | Pregnancy loss |

Through TWAS, we found a correlation between gene expression levels of *TTC38* (Tetratricopeptide Repeat Domain 38) in uterine tissue and pregnancy loss (*Z*=4.83, *P*=1.36×10^−6^; Supplemental Figure S10).

### Gestational Length

The genomic inflations of each sub-cohort GWAS ranged between λ = 0.99 and λ = 1.02, while the GWA meta-analysis across sub-cohorts did not show genomic inflation (Supplemental Figure S3). SNP-based heritability for this trait was 23%. A single SNP, rs144380855, reached genome-wide significance in the meta-analysis of gestational length. rs144380855 appears to be associated with a reduced gestational length (beta = -2.07, *P* = 5.3×10^−10^), and is an intergenic SNP (Table 2). We found evidence of association (*P* < 0.05) in 8 out of 34 SNPs previously reported to be associated with gestational length (Table 3; Supplemental Table S1). These 8 markers were clustered in five genes: *WNT4* (Wnt Family Member 4), *EBF1* (EBF transcription factor 1), *PER3* (Period Circadian Regulator 3), *IL10* (Interleukin 10), and *ADCY5* (Adenalyte Cyclase 5).

**Table 3.**
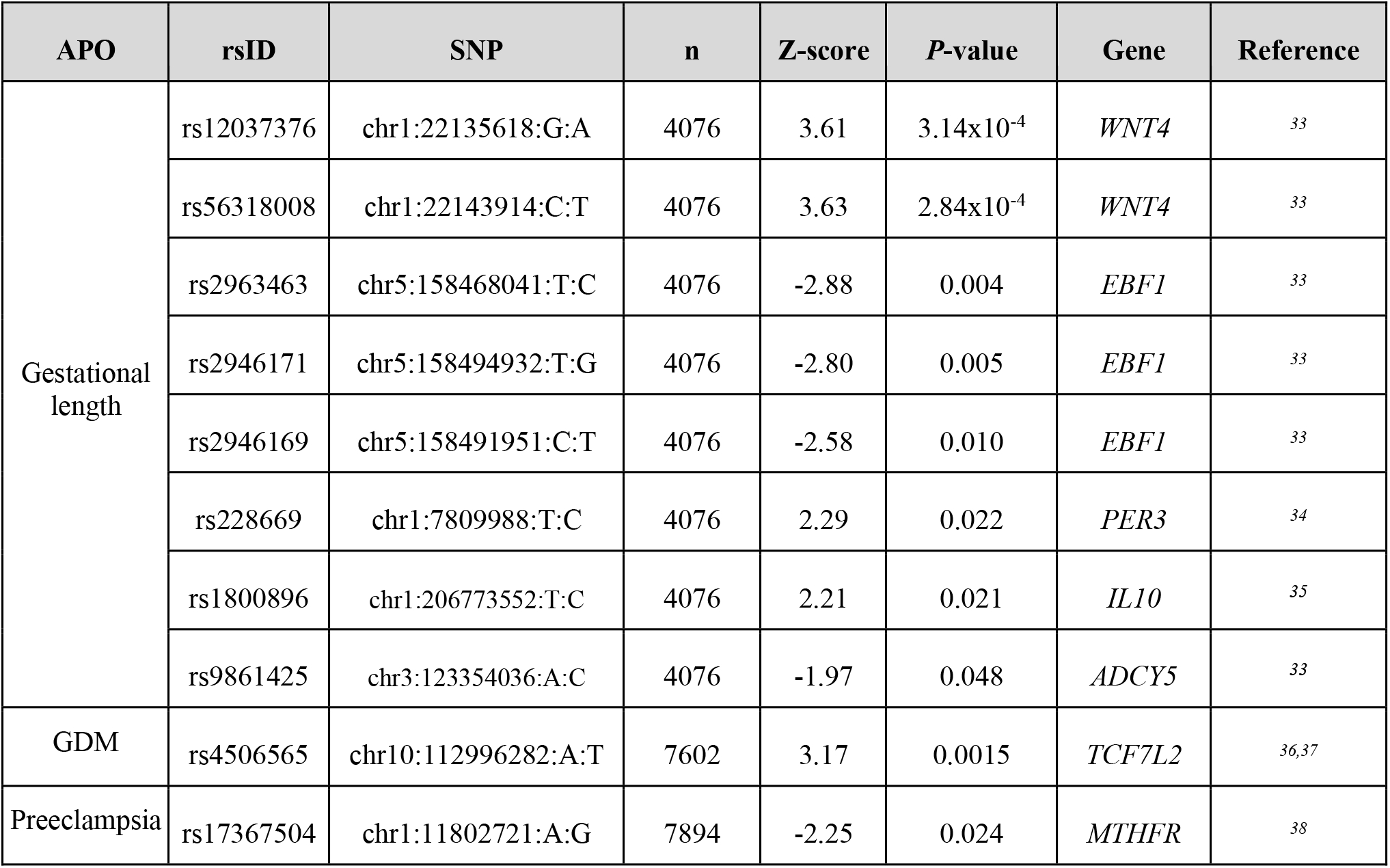
Previously identified variants that were validated in this study (association with relevant APO *P*<0.05). Sample size (n), value of normalized statistic (Z-score) and significance (*P*-value) are shown for the meta-analysis of three sub-populations.

### Gestational Diabetes mellitus

Three significant loci were identified in the GWAS for GDM (Table 2). The top locus included intronic variants rs61167087 and rs72956265, in the gene *ZBTB20*, a Zinc Finger and BTB Domain Containing 20. The second significant association is also an intronic SNP, rs117689036 in *MDGA2*, a MAM Domain-Containing Glycosylphosphatidylinositol Anchor 2. Finally, rs10890563 is a 3’UTR variant on the gene *GUCY1A2*, Guanylate Cyclase 1 Soluble Subunit Alpha 2. Several loci show suggestive associations in the meta-analysis, which we report in the supplemental text (Figure 4; Supplemental Table S2). We did not find genomic inflation in any of the ancestry groups or in the meta-analysis (λ =1.004, Supplemental Figure S5), and we estimated SNP-based heritability to be 0.17.

We also found a positive association consistent with an effect on gestational diabetes for one previously reported variant (rs4506565, *P* = 0.001; Table 3). However, we failed to find any confirmatory signal in 17 other variants (Supplemental Table S1).

### Preeclampsia

We found no SNPs associated with preeclampsia (Supplemental Figure S6). Again, genomic inflation was low (λ =1.005, Supplemental Figure S7), and our SNP-based estimate of heritability (0.02) suggests a weak genetic basis for this APO. We confirmed 1 of 22 previously linked variants: rs17367504 (near gene *MTHFR*) showed a consistent effect across our three studied sub-populations (Table 3; Supplemental Table S1).

## Comment

### Principal Findings

We identified a novel locus associated with gestational length, along with three loci associated with GDM and two additional loci associated with the previously understudied phenotype of pregnancy loss. Our study suggests that variants in *ZBTB20*, a gene that has been previously found to regulate beta cell function and glucose homeostasis in mice^26^, may be involved in the development of GDM. Furthermore, TWAS analysis implicates the gene *TTC38* in pregnancy loss across four tissue types (liver, ovary, pancreas, and uterus). Consistent with this, one of the top SNPs in the pregnancy loss GWAS (rs114058457, Supplementary Table S2), was previously established by Jansen et al. as an eQTL for *TTC38* in blood in a cohort of 4,896 subjects of European ancestry (FDR-corrected *P* < 1.34×10^−5^).^27^

### Clinical Implications

The SNPs that reached genome-wide significance in our study highlight the role of previously unknown genes in our APOs of interest. These genes may further implicate specific biological interactions and pathways that are disrupted in each APO. While not reaching genome-wide significance, we were able to document the directionality of association for several SNPs previously reported for other APOs. These SNPs, now having been discerned by different investigations, are valuable by identifying genes relevant to pathophysiology–which can lead to better prevention and treatment.

Clinical utility of genetic screening is most effective when there is a preventive measure that can be employed. If strong genetic markers for preeclampsia, for instance, had been discovered in pathway genes for mechanisms that could be influenced by aspirin therapy, these would be prime candidates for screening nulliparas who might not have other risk factors that would lead to recommended prophylaxis ^28^. SNPs implicated in shorter gestational length could lead to further screening with cervical length ultrasound or perhaps the administration of preventive progestin therapy. Incorporating genetic screening for APOs into clinical care will necessitate more data and trials, but novel and confirmatory findings such as those presented here are initial steps towards potential clinical utility.

### Research Implications

Given the low heritability computed across all four APOs, it appears that environmental or other factors may play an important role in these phenotypes. One possibility is that individual genetic loci each confer small effects to the phenotype as a whole; indeed, our findings are consistent with this hypothesis. The development of statistical methods that leverage such assumptions, such as polygenic risk scoring, may enable better predictive capacities of models. Evolutionarily, given that variants with a strong effect on infant mortality may face strong negative selection, it may be more likely for rare variants of smaller effect to play a larger role in the genetics of preterm birth^29^. Identifying rare causative variants would require sequencing efforts including whole-genome and whole-exome sequencing, as well as larger study populations of diverse genetic ancestry. Additionally, our study has unearthed potential contributors to preterm birth at multiple levels of granularity—SNP, gene, and tissue. Targeted follow-up functional studies of these layers of genetic factors *in vivo* or *in vitro* may better elucidate the underlying biological mechanisms influencing preterm birth phenotypes and ultimately can shape targeted treatments.

### Strengths and Limitations

While ours is one of the largest GWAS on traits associated with pregnancy (e.g., it is, to our knowledge, the largest of three GWAS on GDM^30,31^), it highlights the limitations of prospective studies in a population of highly diverse genetic ancestry. Namely, this is an imbalanced cohort both in terms of cases-controls and genetic ancestries. Our results would benefit from further validation cohorts and deeper sampling of cases, particularly of individuals of non-European genetic ancestry. Larger sample sizes and future meta-analysis would allow us to explore the diversity of types and etiologies of preterm birth and other APOs. As we noted previously, the environment may play a large role in mediating our APOs, and therefore future directions include conducting gene-by-environment interaction studies to further elucidate the interplay between the two.

### Conclusions

Our study identified novel genetic associations for three pregnancy phenotypes, reporting new variants correlated with gestational length, gestational diabetes, and pregnancy loss. This study confirms the presence of genetic signals in previously identified pathways for preterm birth, GDM, and preeclampsia. Validation of these new associated SNPs in other cohorts will be important in determining their potential clinical usefulness.

## Data Availability

All data produced in the present study will be made available online after peer review, and are available upon reasonable request to the authors

## Acknowledgements

We thank Tatiana Foroud and Kymberleigh Pagel for helpful discussion in the planning stage of this work, and Jelena Radivojac for contributing to the design and logistics of DNA extraction and genotyping.

---

**Figure 1.**
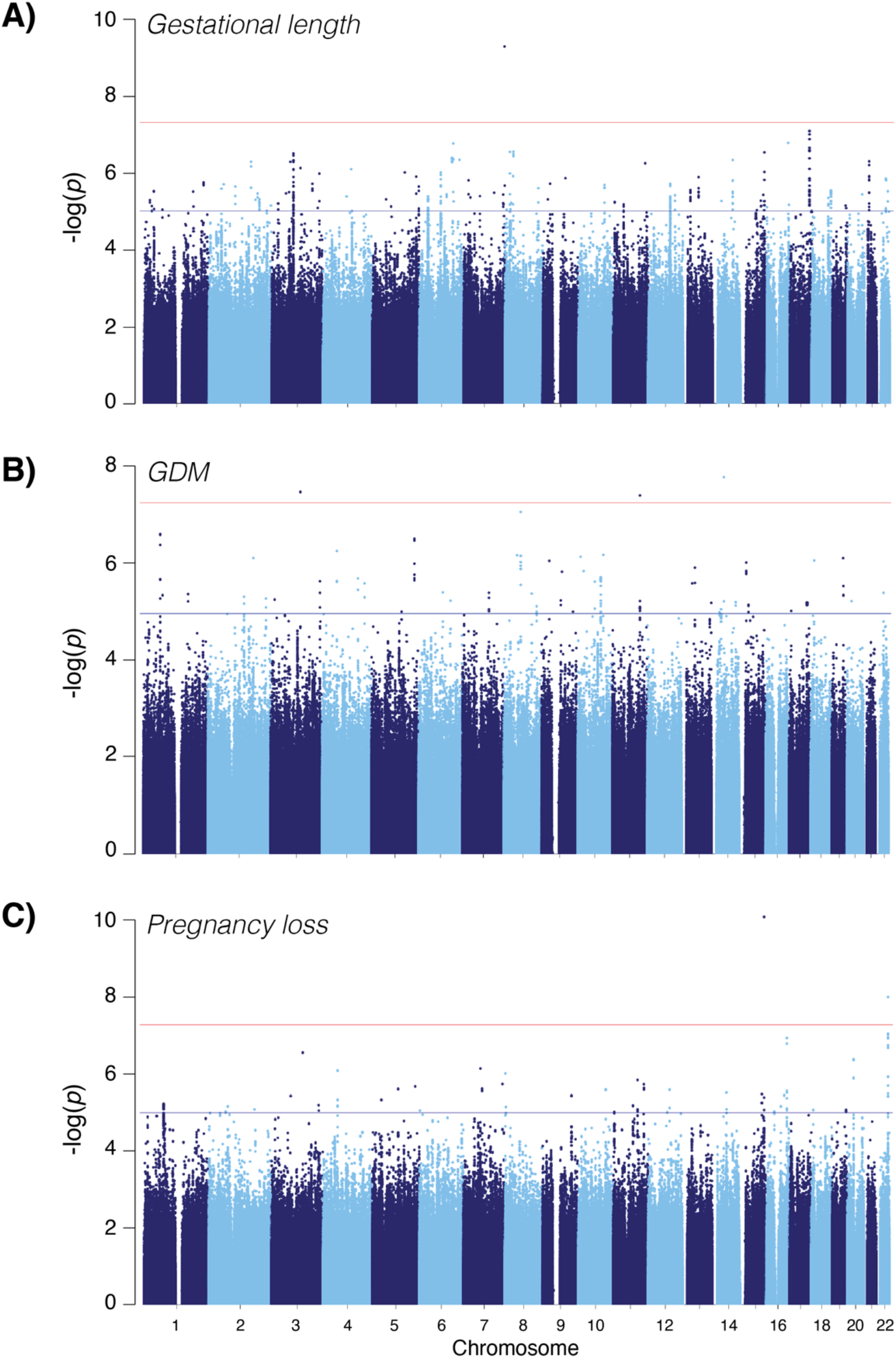
Manhattan Plot of GWAS Results: GWA meta-analysis was carried out using the inverse variance–weighted method implemented in GWAMA. The solid red line indicates the threshold for genome-wide significance (*P* < 5 × 10^−8^), and the blue line represents the threshold for suggestive hits (*P* < 1 × 10^−5^). A) Gestational length; B) GDM; C) Pregnancy loss.

**Figure 2.**
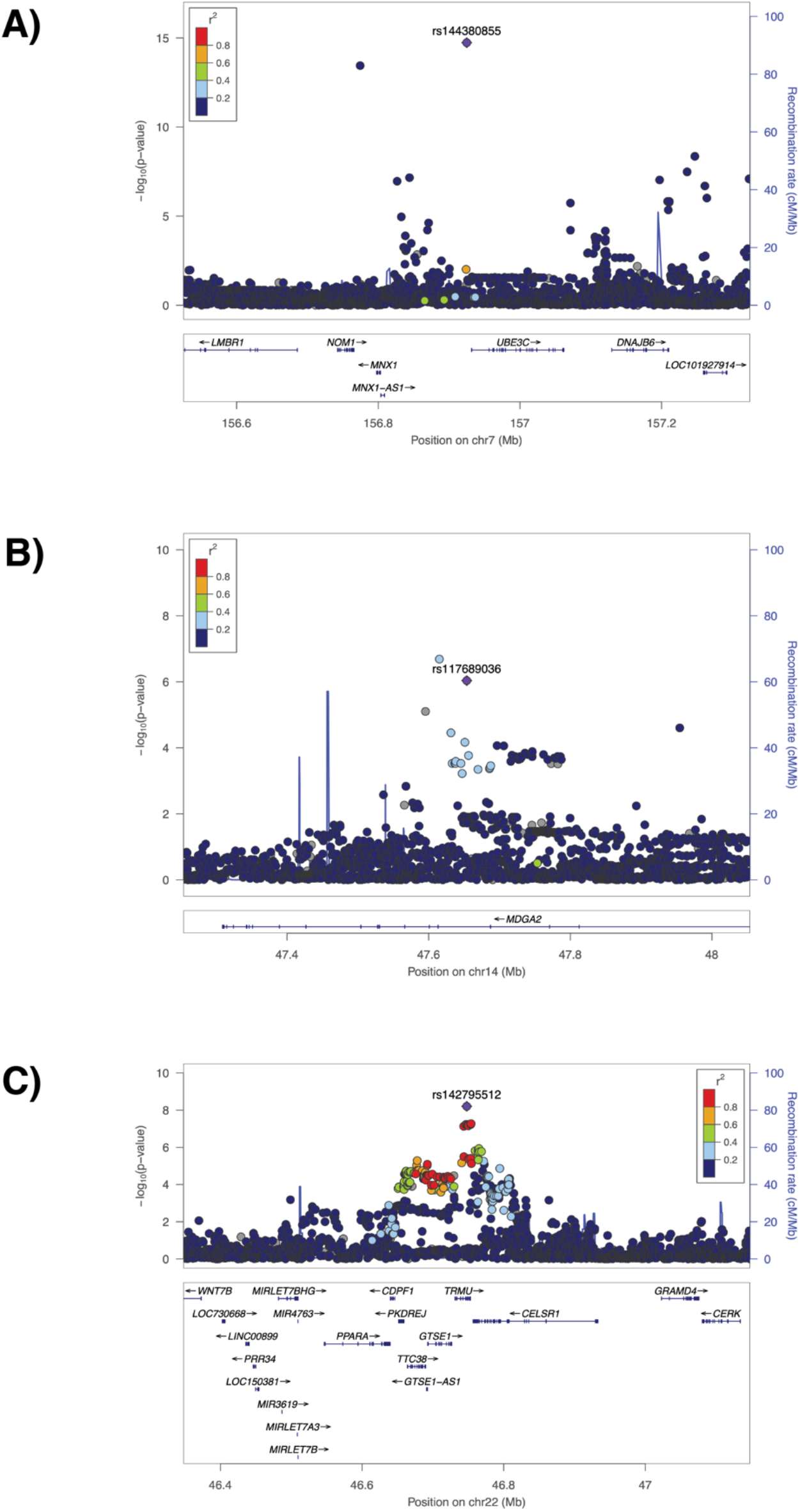
Selective regional association plots demonstrating regional linkage disequilibrium for A) rs144380855 associated with gestational length, B) rs117689036 associated with gestational diabetes, and C) pregnancy-loss associated SNP rs142795512

